# Implementing electronic patient-reported outcome measures in psychiatric urgent care

**DOI:** 10.1101/2025.05.02.25326812

**Authors:** Amber Bailey, Emily Berich-Anastasio, Andrew Ready, Alexander Maclay, Matthew Payne, Robert J. Schloesser

**Affiliations:** Sheppard Pratt, Baltimore, Maryland, USA; University of Maryland, Department of Psychiatry, Baltimore, Maryland, USA

## Abstract

**Background:** Patient-reported outcomes (PRO) have been shown to improve screening and assessment across health care. However, overall implementation of PROs is low in mental health care. Research on PRO implementation in psychiatric urgent care settings is particularly limited.

**Objective:** To analyze barriers and facilitators to the implementation of electronic PROs (ePROs) in psychiatric urgent care clinics.

**Methods:** This study examined ePRO implementation at two Maryland clinics with an average 6,000 patients treated annually. These clinics offer “walk-in” services for patients seeking immediate assessment for mental health conditions and referral to appropriate follow-up care. We used the Learning Evaluation (LE) and RE-AIM frameworks to guide and evaluate the implementation of PROs into administrative and clinical workflows using an ePRO system. Stakeholder feedback informed rapid iteration cycles, driving the development and deployment of technical and procedural modifications. Quantitative data were organized using RE-AIM metrics and analyzed through descriptive statistics and regression analyses. Qualitative data, derived from stakeholder feedback, were analyzed using deductive coding, inductive coding, and sentiment analysis.

**Findings:** 22,610 care episodes were analyzed for the study. Annual ePRO completion increased from 9% in 2021 to 44% in 2023, averaging 63% post-implementation. ePRO completion varied across clinics and was lower among males, Black patients, and those with neurocognitive or substance use disorders, but higher for anxiety, ADHD, and insomnia diagnoses. Adoption increased in 2023, with 17% of care episodes including ePRO data in provider notes, averaging 59% post-implementation. Six iterative modifications were implemented, assisting with ePRO completion and documentation to varying degrees. Qualitative analyses revealed administrative, clinical, and technological factors associated with ePRO implementation and completion rates, as well as an overall positive sentiment towards ePROs.

**Conclusion:** The findings demonstrate the feasibility and sustainability of ePRO implementation in psychiatric urgent care settings.

**Clinical Implications:** Iterative strategies, informed by stakeholder feedback, assist with ePRO implementation in fast-paced clinical environments and can inform future quality improvement efforts for modernization of administrative workflows and clinical practice.

**What is already known on this topic:**

- While research supports patient-reported outcomes (PROs) as effective in mental health treatment when used, multiple barriers have limited their implementation in clinical practice. Currently there is limited knowledge of implementation in psychiatric urgent care settings.

**What this study adds:**

- Using established implementation and evaluation frameworks, we were able to identify and overcome individual and organizational barriers of PRO implementation in psychiatric urgent care.
- We found that continuous communication and feedback can help clinicians and staff with implementation. Iterative modification development based on feedback can improve patient reach and adoption of PROs in clinical practice.

**How this study might affect research, practice or policy:**

- Our findings contribute to gaps in the literature about PRO implementation in psychiatric urgent care and expands on technological and administrative processes that can resolve known barriers to implementation. The methodology used in this study can be modified to numerous real-world healthcare settings for evidence-based quality improvement interventions.

## 1. Background

Patient-reported outcomes (PRO) have demonstrated efficacy in mental health care, contributing to improved outcomes, increased treatment satisfaction, and enhanced clinical decision-making (1). PROs are also effective in screening for depression and anxiety across medical fields (2,3). The value of PROs as a method of quality improvement has been endorsed and facilitated by the American Psychiatric Association (4) and the Joint Commission (5).

Given the benefits of PROs in enhancing mental health care, they would be useful in a healthcare landscape where there is a rising incidence of mental health conditions and increasing emergency department visits (6). Currently, there is limited evidence on PRO implementation in psychiatric emergency, urgent care, or walk-in settings where implementation is challenged by limited evaluation time and high clinical acuity. Concerns persist about how patient populations utilizing emergency services will receive and understand PROs (7). Perceived limited clinical utility (8,9) is an additional barrier. Electronic PROs (ePROs) may resolve some barriers by offering multiple delivery formats, improving time burden, and enabling data integration into electronic medical records (EMRs) (7).

To address barriers and maximize the value of PRO implementation in psychiatric urgent care, utilizing implementation science frameworks can provide systematic methodologies for implementing and evaluating interventions.

The Learning Evaluation framework employs Plan-Do-Study-Act (PDSA) cycles to facilitate continuous quality improvement by collecting and analyzing data, evaluating contextual factors, and generating transferable insights (10).

The RE-AIM framework can complement Learning Evaluation by assessing the intervention’s performance across five key dimensions: Reach, Effectiveness, Adoption, Implementation, and Maintenance. “Reach” is the proportion of the target population engaged by the intervention, while “Effectiveness” measures the intervention’s outcomes. “Adoption” measures the level of organizational participation, “Implementation” assesses the intervention’s cost and scalability, and “Maintenance” examines the intervention’s sustainability over time. By focusing on real-world applicability, RE-AIM is particularly useful for addressing the contextual challenges specific to different industries and organizations (11).

This study used the Learning Evaluation and RE-AIM frameworks to develop and evaluate an implementation process for an ePRO intervention in psychiatric urgent care.

### 1.1 Objective

The primary objective was to identify the barriers and facilitators influencing implementation. The secondary objective was to rapidly develop modifications that resolve barriers and enhance facilitators in real-world clinical settings.

## 2. Methods

### 2.1 Setting

From August 2021 to December 2023, an electronic patient-reported outcomes (ePRO) intervention was implemented at two psychiatric urgent care (PUC) clinics operated by Sheppard Pratt, a large private, nonprofit mental health provider in the United States. Located in central Maryland, the two clinics—Location B (Howard County) and Location T (Baltimore County)—serve around 6,000 patients annually. Location B was newly established before the study period, while Location T has been operational for many years. Both clinics offer walk-in psychiatric assessments and coordinate referrals through multidisciplinary clinical teams. Patients aged five and older were eligible for evaluation and participation in the ePRO intervention.

Oversight was provided by an ePRO implementation team, supported by a digital services team (Supplementary Table 1). Feedback was collected from urgent care clinical and administrative managers (Supplementary Table 2).

### 2.2 Intervention

Supplementary Figure 1 illustrates the workflow of the ePRO intervention (12). A custom health IT application (see Section 2.4 Technology) was used to collect, score, and transfer ePRO results into the EMR. Notifications triggered by HL7 registration and scheduling events were sent via email or SMS/text; on-demand notifications were also available via QR codes or direct links (Supplementary Figure 2a). Dashboards enabled real-time monitoring of ePRO deployment and completion by staff, accessible through the EMR or health system workstations. Spanish-language options were available through an on-screen selector (Supplementary Figure 2b).

Completed ePROs generated score reports transmitted to the EMR within three minutes, viewable by clinicians for integration into service notes. Item-level data were available on the ePRO dashboard. Implementation “Reach” was measured as the proportion of admitted patients with completed ePROs. “Adoption” was measured as the proportion of completed ePROs documented in service notes.

Patients aged 12 and older completed ePROs independently; parent-proxy reports were used for younger children. The battery included PROMIS measures and screeners for alcohol use, tobacco use, elder abuse, and fall risk, selected collaboratively by the ePRO team and clinical managers (Supplementary Table 3).

### 2.3 Implementation Frameworks and Metrics

Implementation followed the Learning Evaluation (LE) framework, with RE-AIM guiding outcome evaluation. Contextual factors influencing implementation were captured during governance meetings and informed iterative refinements using Plan-Do-Study-Act (PDSA) cycles.

Weekly governance meetings for the ePRO implementation team reviewed stakeholder input and RE-AIM metrics, provided from monthly site meetings with urgent care staff. Supplementary Figure 3 depicts the governance process for identifying barriers, setting goals, and tracking progress (13).

Intervention modifications were guided by SMART criteria (14) and documented using the Framework for Reporting Adaptations and Modifications-Enhanced (FRAME) (15). During the “act” phase of PDSA, modifications were further expanded into descriptions of end-user needs. In the “plan” phase, itemized tasks for the modifications were refined during discovery sessions. The “do” phase involved completing these tasks during two-week sprint cycles.

### 2.4 Technology

The ePRO system integrated open-source tools with Sheppard Pratt’s electronic health record (EHR). Research Electronic Data Capture (REDCap) managed assessments, scoring, support tickets, transcripts, and system modifications via a secure, web-based interface. It supported audit trails, exports, and external integration (16,17).

R supported statistical analysis and data visualization, while R Shiny enabled interactive dashboards. Drupal managed user access and support ticket collection. Docker images hosted submodules including a Node.js HL7 ingestion client and R Shiny dashboards. Components were linked via MySQL databases and Python scripts for automated ePRO deployment. HL7 interfaces enabled real-time event-based deployment, with custom document-based integration for EMR reporting.

### 2.5 Data Sources

Demographic, registration, and scheduling data were drawn from the revenue cycle system (AthenaIDX, AthenaHealth Inc., Boston, MA). Diagnostic data came from the EMR (Sunrise, Altera Digital Health Inc., San Jose, CA). A licensed mental health clinician and a board-certified psychiatrist grouped International Classification of Diseases (ICD) codes into DSM-5–based clinical categories (Supplementary Table 4).

Additional data sources included system logs (ePRO “Reach” and “Adoption”) and stakeholder feedback from governance meetings and support tickets. All feedback and documentation were stored in a database used during the PDSA “study” phase to ensure transparency and accountability.

### 2.6 Statistical Methods

A mixed-methods approach evaluated “Reach,” “Adoption,” “Implementation,” and “Maintenance” (Supplementary Table 5). Quantitative analyses included descriptive statistics and logistic regression, with interpretations based on effect sizes (18). Patients identifying as “Other” sex were excluded from regression due to small sample size (n = 6). Each “care episode” represented a distinct admission period at a clinic.

Qualitative data were analyzed using deductive and inductive thematic coding of stakeholder feedback, categorized by date, stakeholder type, and implementation themes (19). Sentiment analysis was conducted using the Sentimentr package to assess sentence-level polarity (20). Additional analyses included Welch’s T-Test and ANOVA.

All patient data were de-identified prior to analysis. No patients were excluded from completing ePROs. For qualitative analyses, a “patient” was defined as any individual receiving treatment.

## 3. Results

### 3.1 Quantitative Analysis

This analysis assessed ePRO completion (“Reach”), ePRO documentation (“Adoption”), feedback and modification processes (“Implementation”), and the sustainability of these metrics over time (“Maintenance”).

#### 3.1.1 Reach

Among 22,610 care episodes, 5,383 completed ePROs (24%). Patients averaged 1.3 admissions (*SD* = 0.76). Completion rates significantly increased over time (*R*² = 0.83, *p* = < .0001), peaking at 73% (Figure 1). ePROs averaged 50 seconds to complete (SD = 18.1). Most patients were adults aged 18–64 (57%), female (57%), and White (53%), with a mean age of 28 (SD = 17) (Table 1a). Location T had more admissions (64%) than Location B (36%) but 27% decreased odds of ePRO completion (Table 1a). This disparity was consistent across age groups, with adult, geriatric, pediatric, and proxy-report care episodes at Location T all showing significantly reduced odds of completion compared to Location B (Table 1b).

**Figure 1.**
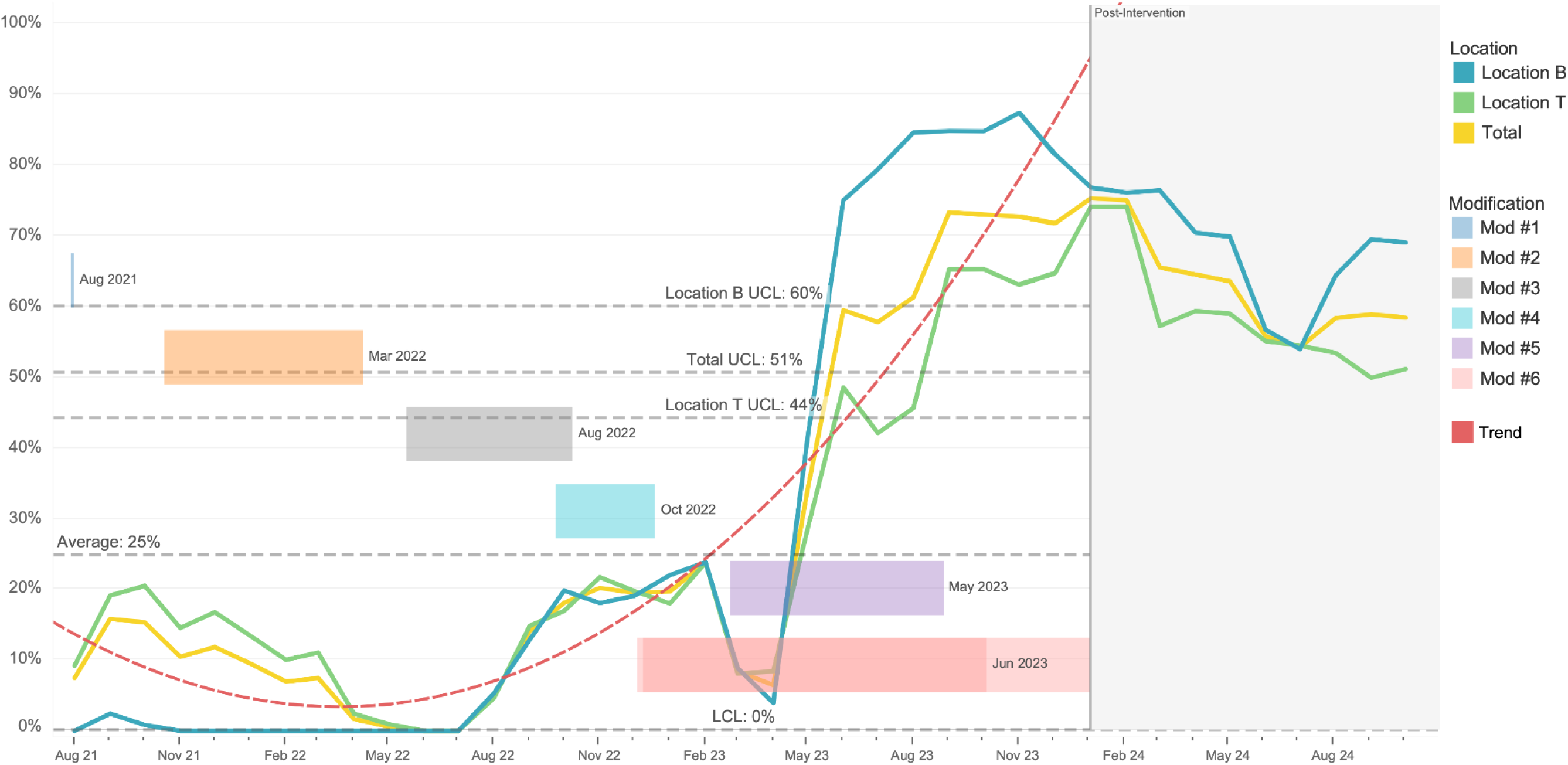
Monthly ePRO completion with trendline Figure 1 displays ePRO completion (“Reach”) throughout the implementation period (August 2021 - December 2023) and post-implementation (January - October 2024 highlighted in gray). A total of 5,383 patients completed ePROs during the intervention period. The equation for the polynomial trend line (dotted red line) during the implementation period is *y = 2.49e-10x^3^ + −3.14e-05x^2^ + 1.31x+ −18218.6* (*R^2^* = 0.83, *P* = <0.0001). Upper (UCL) and lower (LCL) control limits for ePRO completion during the implementation period are displayed, stratified by location. Both locations had a LCL of 0% but different UCLs: 60% for Location B, 44% for Location T, and 51% for both locations combined (Total). The average monthly ePRO completion for the implementation period was 25%. Post-implementation average was 63%. A gantt chart of the 6 modifications implemented displayed (descriptions in Supplementary Table 6). The length of each gantt bar represents the amount of time needed to complete the modification. The month and year presented to the right of each gant bar represents when the modification was completed. Mod #6 has two completion dates because the modification was completed at two different times for Location B and Location T.

**Table 1a.** Comparing patient demographics and ePRO completion during implementation period (August 2021 - December 2023)

| Variable | Summary Statistics |  |  | ePRO Completion Regression |  |  |
| --- | --- | --- | --- | --- | --- | --- |
|  | Total, N = 22,610 | ePRO incomplete, N = 17,221 | ePRO complete, N = 5,383 | log(OR) (95% CI) | log(OR) Interpretation | p-value |
| **Location** |  |  |  |  |  |  |
| Location B | 8,171 (36%) | 5,929 (73%) | 2,237 (27%) | — |  | — |
| Location T | 14,439 (64%) | 11,292 (78%) | 3,146 (22%) | -0.31 (-0.38, -0.25) | 27% ↓ | <0.001 |
| **Sex** |  |  |  |  |  |  |
| Female | 12,986 (57%) | 9,804 (75%) | 3,182 (25%) | — |  | — |
| Male | 9,618 (43%) | 7,417 (77%) | 2,201 (23%) | -0.10 (-0.17, -0.04) | 10% ↓ | 0.001 |
| Other | 6 (<0.1%) |  |  | NA | NA | NA |
| **Age** | 28 (17) | 28 (17) | 27 (16) | — |  | — |
| **Age Range** |  |  |  |  |  |  |
| Adult 18-64 yrs | 12,859 (57%) | 9,723 (76%) | 3,131 (24%) | — |  | — |
| Pediatric 12-17 yrs | 6,702 (30%) | 5,137 (77%) | 1,564 (23%) | -0.07 (-0.14, 0.00) | 7% ↓ | 0.052 |
| Proxy-Report 5-11 yrs | 2,136 (9.4%) | 1,616 (76%) | 520 (24%) | 0.04 (-0.07, 0.15) | 4% ↑ | 0.5 |
| Geriatric 65+ yrs | 913 (4.0%) | 745 (82%) | 168 (18%) | -0.38 (-0.55, -0.21) | 32% ↓ | <0.001 |
| **Race** |  |  |  |  |  |  |
| White | 12,060 (53%) | 9,031 (75%) | 3,024 (25%) | — |  | — |
| Black/African-American | 7,646 (34%) | 5,938 (78%) | 1,708 (22%) | -0.16 (-0.22, -0.09) | 15% ↓ | <0.001 |
| Other | 2,261 (10%) | 1,754 (78%) | 506 (22%) | -0.20 (-0.31, -0.10) | 18% ↓ | <0.001 |
| Asian | 643 (2.8%) | 498 (77%) | 145 (23%) | -0.22 (-0.42, -0.03) | 20% ↓ | 0.023 |

**Table 1b.**
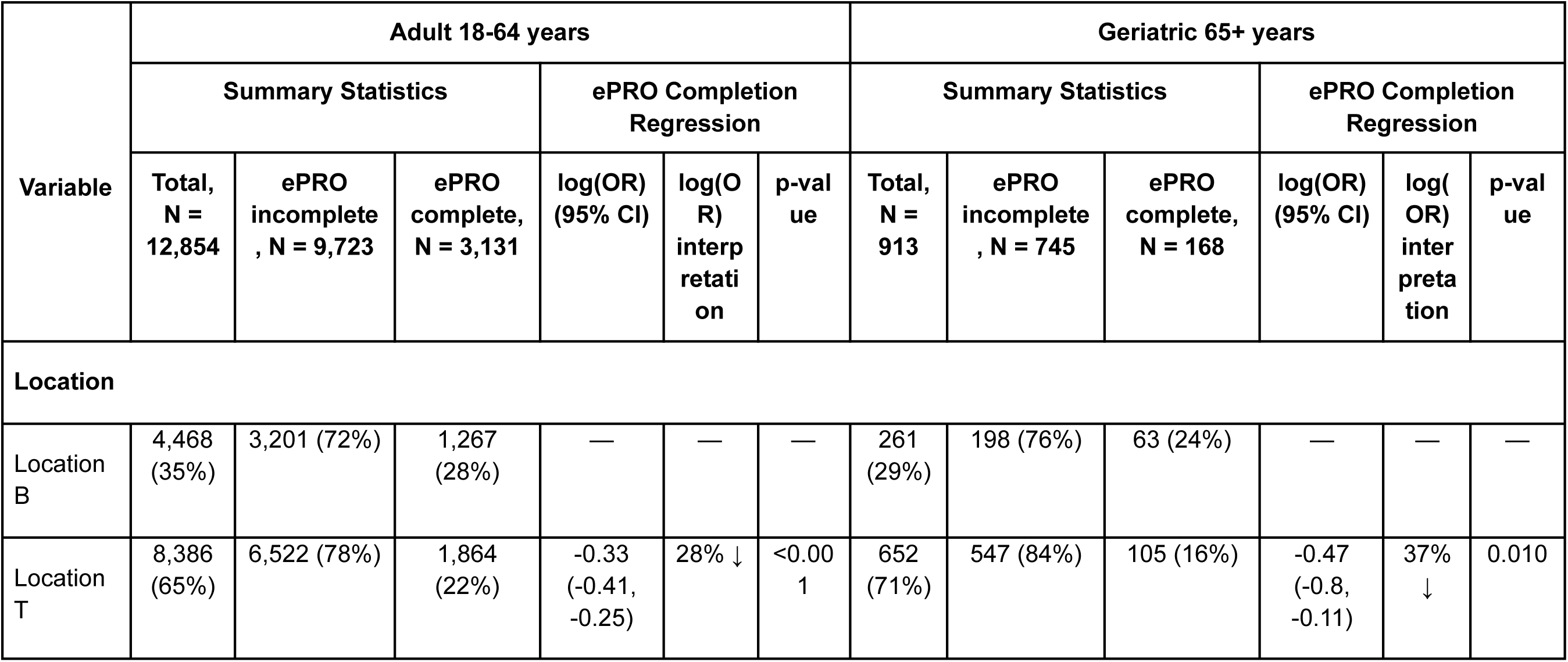
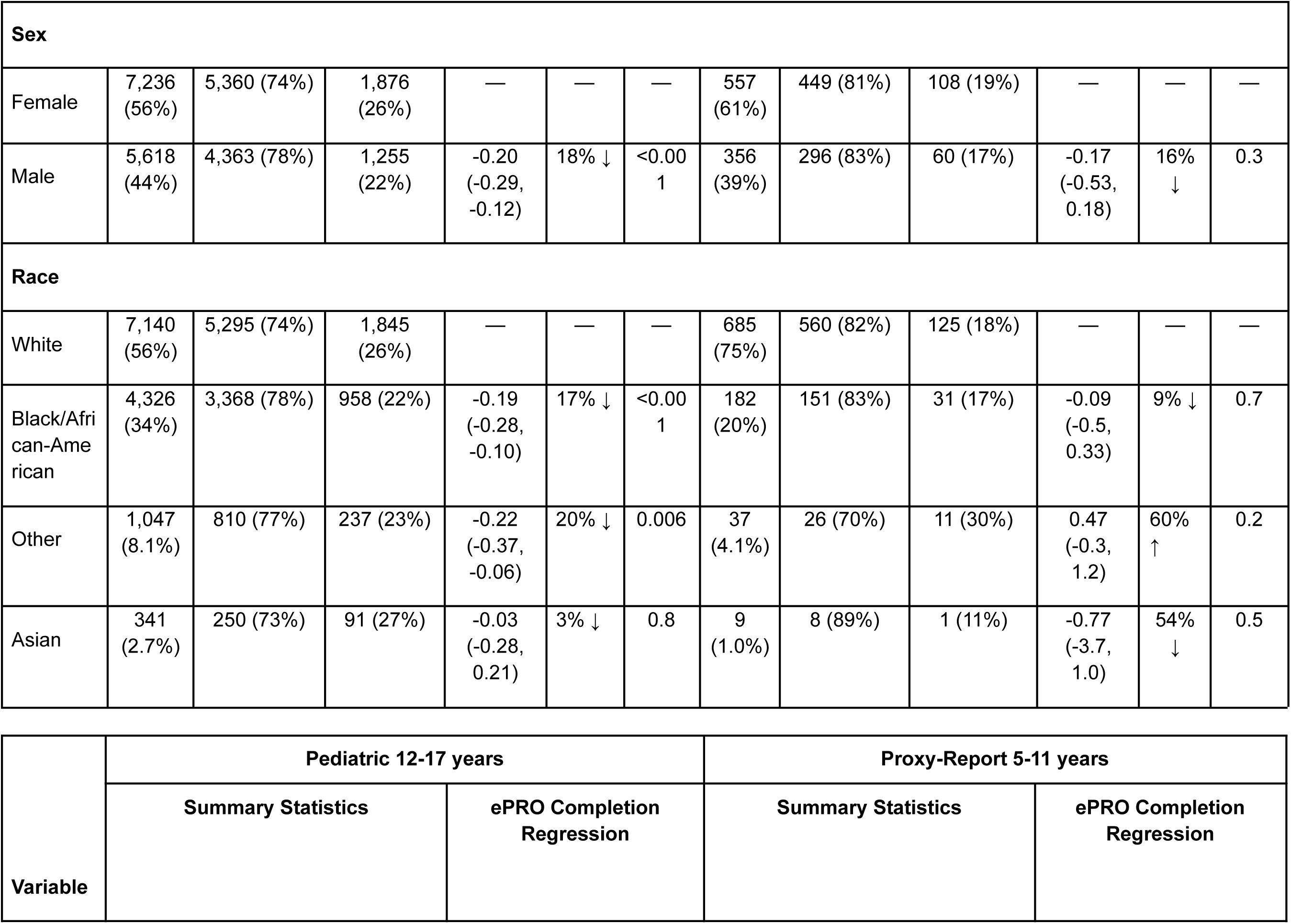
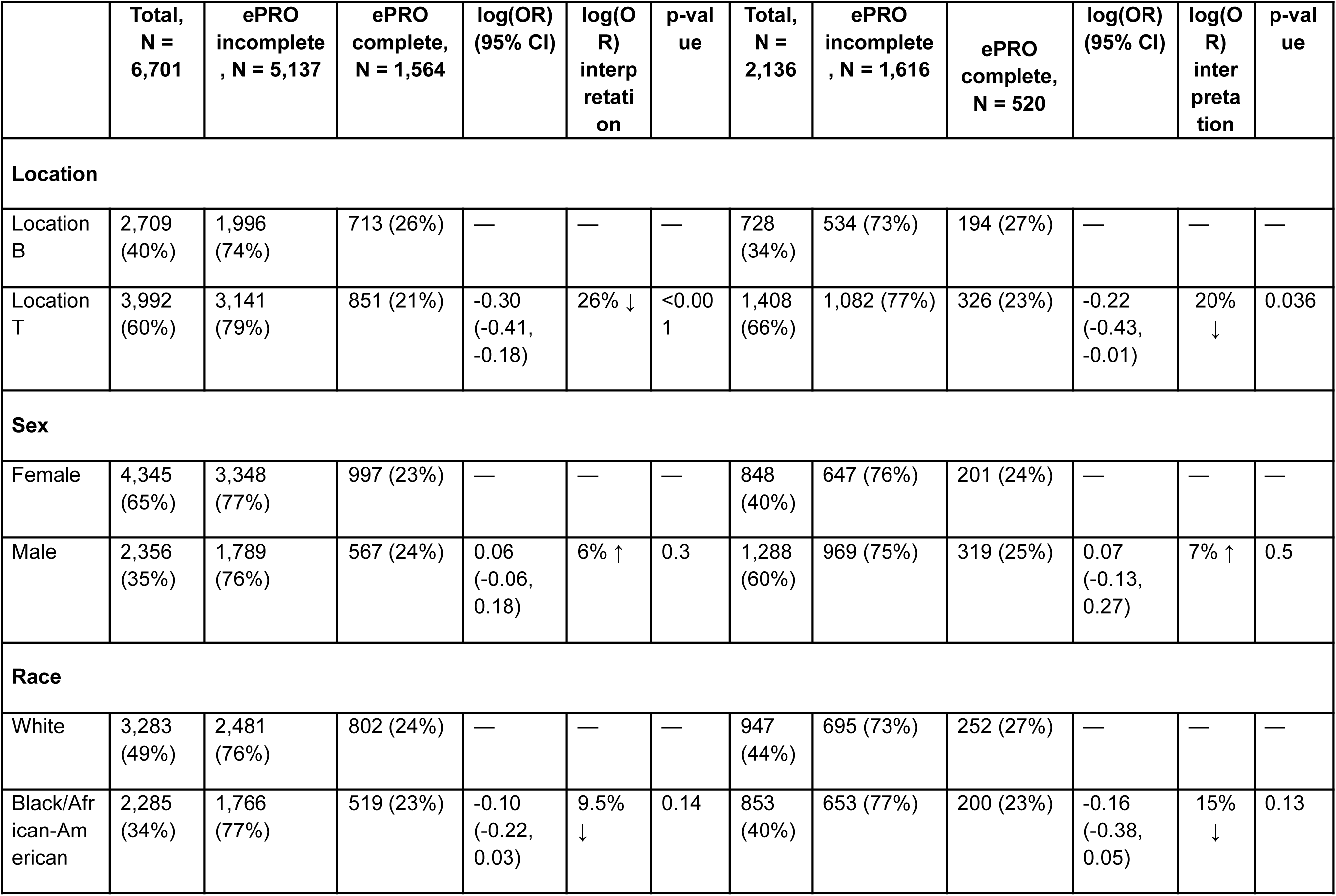
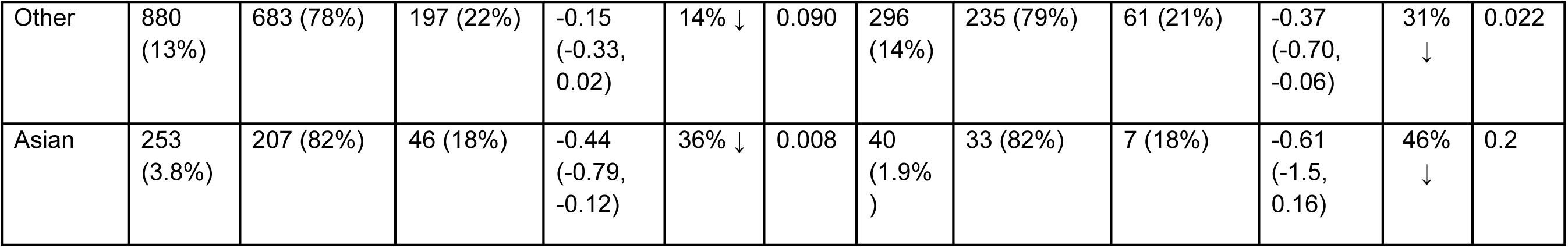
Age group stratification of ePRO completion Tables 1a and 1b display a summary (“Summary Statistics”) and logistic regression ePRO completion (“ePRO Completion”) for urgent care patients by demographic variable (3a) and age group (3b). “Summary Statistics” displays the count of all care episodes admitted and stratified by demographic variable (“Total”), the count of care episodes who did not complete an ePRO during their admission (“ePRO incomplete”), and the count of care episodes who completed an ePRO during their admission (“ePRO complete”). For the “Total” column, all variable statistics are presented as n (%) for the column. For “Age”, the data is presented as Mean (SD). “ePRO incomplete” and “ePRO complete” statistics are presented as n (%) for the row (i.e. variable). “ePRO Completion Regression” displays the logistic regression (“log(OR)”) of completing ePROs, the associated confidence interval (“95% CI”), and *p* (“p-value”). The logistic regression and confidence interval are not exponentiated. “log(OR) interpretation” is calculated as (1 − 𝑒^𝑙𝑜𝑔(𝑂𝑅)^) × 100 . Values are rounded to the nearest zero. The absolute value was used for negative values. Down arrows ↓ indicate decreased odds and Up arrows ↑ indicate increased odds. NA indicates the variable was not included in the logistic regression.

Adults identifying as male, Black, or Other race had 18%, 17%, and 20% decreased odds of completing ePROs, respectively, compared to patients identifying as female or White (Table 1b). Pediatric patients identifying as Asian had 36% decreased odds and proxy patients identifying as Other race had 31% decreased odds of completion compared to patients identifying as White (Table 1b).

A total of 54,302 diagnoses were recorded (mean = 2.2 per episode, *SD* = 1.1) in the EMR for the admitted care episodes. The most frequent categories included depressive, anxiety, and alcohol/substance use disorders (Table 2). Decreased odds of completion were demonstrated in patients with neurocognitive, psychotic, or alcohol/substance use diagnoses, and higher in patients with with ADHD, anxiety, autism spectrum disorder (ASD), OCD, cluster B personality disorders, insomnia, and when no diagnosis was listed (Table 2).

**Table 2.** Comparing diagnostic categories and ePRO completion during implementation period Table 2 displays the (“Summary Statistics”) and logistic regression ePRO completion (“ePRO Completion”) of diagnostic category entries in the EMR for patients admitted to urgent care during the implementation period. “Summary Statistics” displays the count of all diagnostic category entries (“Total”), the count of diagnostic categories that did not complete an ePRO during admission (“ePRO incomplete”), and the count of diagnostic categories that completed an ePRO during admission (“ePRO complete”). For the “Total” column, all diagnostic category statistics are presented as n (%) for the column. The “Total” is higher than the actual number of admitted patients (n = 22,610) because most patients had more than one diagnostic category assigned in the EMR; there was an average of 2.2 diagnostic categories per patient (*SD* = 1.1). “ePRO incomplete” and “ePRO complete” statistics are presented as n (%) for the row (i.e. diagnostic category). “ePRO Completion Regression” displays the logistic regression (“log(OR)”) of completing ePROs, the associated confidence interval (“95% CI”), and *p* (“p-value”). The logistic regression and confidence interval are not exponentiated. “log(OR) interpretation” is calculated as (1 − 𝑒^𝑙𝑜𝑔(𝑂𝑅)^) × 100 . Values are rounded to the nearest zero. The absolute value was used for negative values. Down arrows ↓ indicate decreased odds and Up arrows ↑ indicate increased odds.

| Diagnostic Category | Summary Statistics |  |  | ePRO Completion Regression |  |  |
| --- | --- | --- | --- | --- | --- | --- |
|  | Total, N = 54,302 | ePRO incomplete, N = 40,986 | ePRO complete, N = 13,316 | log(OR) (95% CI) | log(OR) interpretation | p-value |
| All Diagnostic Categories | 54,302 (100%) | 40,986 (68%) | 13,316 (32%) | — | — | — |
| Depressive Disorders | 14,979 (28%) | 11,196 (75%) | 3,783 (25%) | 0.04 (0.00, 0.08) | 4% ↑ | 0.055 |
| Anxiety Disorders | 9,841 (18%) | 7,126 (72%) | 2,715 (28%) | 0.16 (0.11, 0.21) | 17% ↑ | <0.001 |
| Alcohol and Substance Use Disorders | 6,165 (11%) | 4,875 (79%) | 1,290 (21%) | -0.20 (-0.27, -0.14) | 18% ↓ | <0.001 |
| Bipolar Disorders | 4,307 (7.9%) | 3,278 (76%) | 1,029 (24%) | -0.03 (-0.11, -0.04) | 3% ↓ | 0.4 |
| Psychosis, Schizophrenia Spectrum, Delusional Disorders | 3,618 (6.7%) | 2,966 (82%) | 652 (18%) | -0.39 (-0.48, -0.30) | 32% ↓ | <0.001 |
| ADHD | 3,427 (6.3%) | 2,498 (73%) | 929 (27%) | 0.14 (0.06, 0.21) | 15% ↑ | <0.001 |
| Trauma Disorders | 3,255 (6.0%) | 2,485 (76%) | 770 (24%) | -0.05 (-0.13, 0.04) | 5% ↓ | 0.3 |
| Adjustment Disorders | 1,509 (2.8%) | 1,167 (77%) | 342 (23%) | -0.10 (-0.22, 0.02) | 9% ↓ | 0.1 |
| Severe and Treatment Resistant Mood Disorders | 1,269 (2.3%) | 988 (78%) | 281 (22%) | -0.13 (-0.27, 0.00) | 12% ↓ | 0.055 |
| Impulse Control Disorders | 1,216 (2.2%) | 921 (76%) | 295 (24%) | -0.01 (-0.15, 0.12) | 1% ↓ | 0.9 |
| Autism Spectrum Disorder (ASD) | 874<br>(1.6%) | 635 (73%) | 239 (27%) | 0.15 (0.00, 0.30) | 16% ↑ | 0.052 |
| OCD | 816<br>(1.5%) | 581 (71%) | 235 (29%) | 0.22 (0.07, 0.37) | 25% ↑ | 0.005 |
| Cluster B Personality Disorders | 745<br>(1.4%) | 536 (72%) | 209 (28%) | 0.18 (0.02, 0.34) | 20% ↑ | 0.025 |
| Eating Disorders | 557<br>(1.0%) | 428 (77%) | 129 (23%) | -0.07<br>(-0.27, 0.12) | 7% ↓ | 0.5 |
| Neurodevelopmental Disorders, Other | 360<br>(0.7%) | 286 (79%) | 74 (21%) | -0.23<br>(-0.49, 0.02) | 21% ↓ | 0.084 |
| Neurocognitive Disorders | 298<br>(0.5%) | 243 (82%) | 55 (18%) | -0.36<br>(-0.66, -0.07) | 30% ↓ | 0.016 |
| No Dx Listed | 287<br>(0.5%) | 200 (70%) | 87 (30%) | 0.29 (0.04, 0.54) | 34% ↑ | 0.023 |
| Gender Dysphoria | 233<br>(0.4%) | 182 (78%) | 51 (22%) | -0.15<br>(-0.47, 0.16) | 14% ↓ | 0.4 |
| DMDD | 206<br>(0.4%) | 150 (73%) | 56 (27%) | 0.14<br>(-0.17, 0.44) | 15% ↑ | 0.4 |
| Somatic Disorders | 171<br>(0.3%) | 133 (78%) | 38 (22%) | -0.13<br>(-0.50, 0.22) | 12% ↓ | 0.5 |
| Insomnia | 169<br>(0.3%) | 112 (66%) | 57 (34%) | 0.45 (0.12, 0.77) | 57% ↑ | 0.006 |

#### 3.1.2 Adoption

In 2023, 17% (700/4,068) of care episodes had ePROs documented in the EMR, increasing significantly over time (*R*² = 0.80, *p* = < .0001) (Figure 2). Documentation was completed by 22 of 60 providers: 11 nurse practitioners (629 care episodes), 6 psychiatry residents (20), 3 psychiatrists (34), 1 licensed clinical professional counselor (LCPC) (20), and 1 licensed clinical social worker (LCSW) (7). No documentation occurred in 2021; 2022 had only 20 records.

**Figure 2.**
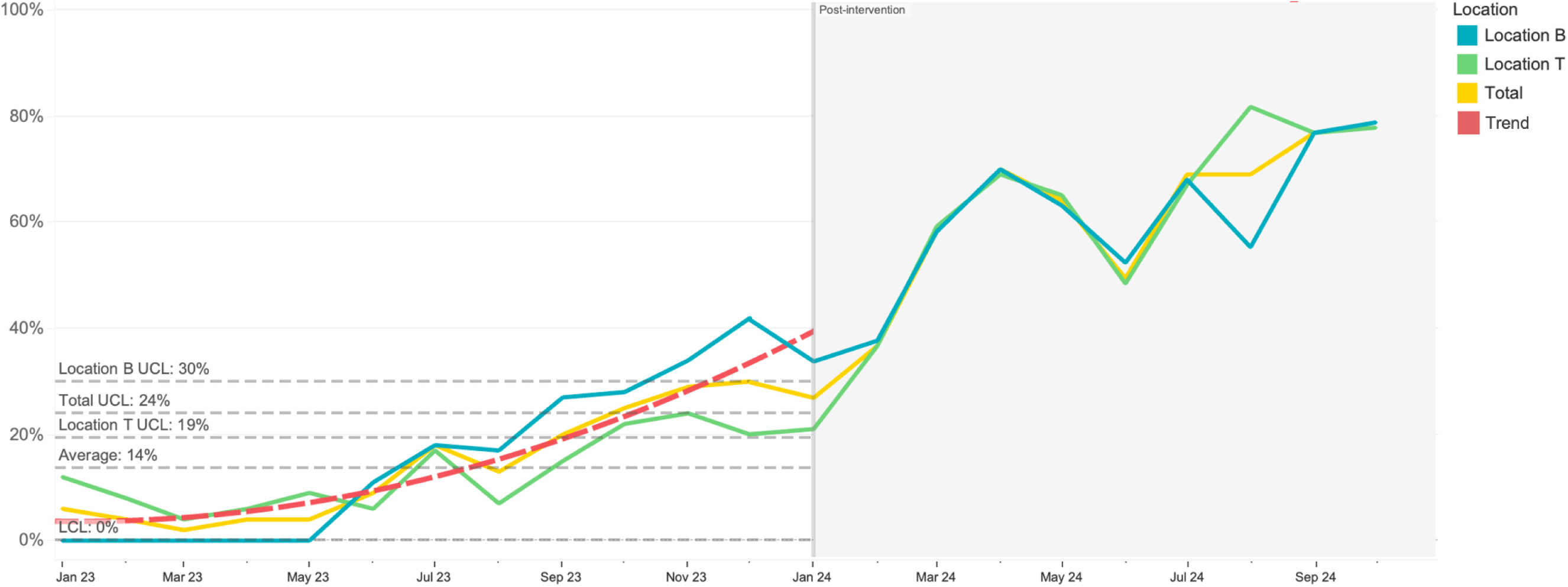
Monthly ePRO documentation in 2023 Figure 2 shows ePRO documentation (“Adoption”) throughout 2023, the final year of the implementation period (2023) and post-implementation (January - October 2024 highlighted in gray). The equation for the polynomial trend line (dotted red line) during the implementation period is *y = 2.80969e-06x^2^ + −0.252506x + 5673.2* (*R^2^* = 0.801437, *P* = < 0.0001). ePRO documentation is defined as the process of incorporating ePRO data into an EHR appointment note. Upper (UCL) and lower (LCL) control limits for the intervention period are displayed, stratified by location. Both locations had a LCL of 0% but different UCLs: 30% for Location B, 19% for Location T, and 24% for both locations combined (Total). The average ePRO documentation for the implementation period was 14%. Post-implementation average was 59%.

#### 3.1.3 Implementation

Implementation efforts included 10 site governance meetings, 2 training sessions, 2 on-site observations, and 1 provider 1:1 meeting—totaling 9.5 hours of engagement with the urgent care team. Five support tickets were submitted, and six system modifications were implemented to enhance workflow and usability (Figure 1, Supplementary Table 6).

#### 3.1.4 Maintenance

Post-implementation, ePRO completion averaged 63% (*SD* = 7%) (Figure 1). Documentation rose to an average of 59% (*SD* = 17%) (Figure 2), completed by 34 of 41 providers, including 18 nurse practitioners (1,972 care episodes), 6 LCSWs (317), 6 psychiatry residents (50), 2 psychiatrists (27), 1 fellow (2), and 1 LCPC (124).

### 3.2 Qualitative Analysis

Analysis of 53 feedback comments and audit notes (137 sentences) identified six key themes: administrative processes, clinical adoption, time, technology, training, and accessibility. Barriers were noted more frequently (32 comments) than facilitators (21 comments), with administrative process (28 comments) and clinical adoption being most common (12 comments). Supplementary Table 7 provides examples of these themes.

#### 3.2.1 Sentiment

Sentiment scores were slightly positive (mean = 0.10, SD = 0.24), increasing to 0.13 after adjustment. Of 137 sentences, 26 were negative, 37 neutral, and 74 positive. The lowest sentiment (−0.56) reflected clinical utility concerns (Oct 2021); the highest (0.82) praised workflow adaptations (June 2023). Facilitator comments had more positive sentiment (mean = 0.18, *SD* = 0.22) than barriers (mean = 0.04, *SD* = 0.24), *t*(120) = 3.45, *p* = .001. Technology had the lowest mean sentiment (−0.08, *SD* = 0.14), and administrative processes the highest (0.12, *SD* = 0.22). No significant differences were found among the comment themes or date.

#### 3.2.2 Reach

Low completion at the start of implementation for Location B was linked to its recent opening in 2021. COVID-19 protocols allowing in-person visits in 2022 led to a completion decline as staff were not informing patients about ePRO emails or providing tablets (Figure 1). Training in August 2022 improved rates, though communication gaps (e.g., instructing patients about assistance and Spanish translation availability) and tech issues (e.g., March 2023 SMS outage) temporarily hindered completion (Figure 1). After system fixes, rates rebounded to 59% by June 2023. The introduction of ”quick registration” in October 2022 streamlined admissions, reducing ePRO notification delays to 3-5 minutes (Mod #6 in Figure 1).

#### 3.2.3 Adoption

Low documentation in 2021 and 2022 was tied to concerns over clinical utility and documentation burden. Integration of the ePRO system with Centers for Medicare & Medicaid Services (CMS) Quality Payment Program (QPP metrics (21) (Supplementary Table 8) led to improved clinical alignment and reduced provider burden, increasing documentation rates (Figure 2). In 2024, clinical leads requested reports on clinician-level ePRO documentation.

#### 3.2.4 Implementation

Efficient processes (e.g., “quick registration”) and clinician engagement facilitated implementation. Location T’s initial concerns about patient burden with “quick registration” processes was overcome by observing Location B’s success, leading to eventual adoption (Mod #6 in Figure 1). Aligning ePROs with CMS QPP metrics increased clinician interest, reducing documentation burden in the EMR (Figure 2, Mod #4 in Figure 1). Technology enhancements like dashboard filtering and SMS increased ePRO access (Mod #3 and Mod #5 in Figure 1).

#### 3.2.5 Maintenance

Routine check-ins by lead administrators to ensure administrative workflows, updated training materials for staff, and efficient registration processes helped maintain strong post-implementation performance (Figure 1 and Figure 2). Despite challenges (staffing fluctuations, inconsistent Wi-Fi connectivity, patient technical literacy), completion rates remained above 50%, and staff confidence in workflows remained high.

## 4. Discussion

This study evaluated the implementation of ePROs in psychiatric urgent care using the Learning Evaluation framework and assessed outcomes through the RE-AIM model. Findings support the feasibility of ePRO integration in this setting and underscore the value of iterative feedback in identifying key barriers and facilitators. To our knowledge, this is the largest reported ePRO implementation in psychiatric urgent care, expanding on prior, smaller studies in inpatient and outpatient mental health contexts (22,23).

### 4.1 ePRO Completion and Patient Demographics

Completion rates varied by demographic and operational factors. Adults aged 18–64, White patients, and females were more likely to complete ePROs, consistent with prior research (24). However, unlike previous findings, we observed no significant disparities in completion among pediatric patients (25). This may be due to increased smartphone access among adolescents (26) and the successful use of SMS/text reminders—accounting for over 50% of completions in 2023.

Lower completion at Location T may reflect differences in operational maturity and greater change management challenges compared to Location B, which benefited from fewer legacy workflows and faster adoption of innovations like “quick registration.” These findings echo literature indicating that established clinical practices may be resistant to change, emphasizing the need for site-specific implementation strategies (27).

### 4.2 ePRO Completion and Diagnosis

Completion rates also varied by diagnosis. Patients with neurocognitive disorders, psychosis, schizophrenia spectrum, and substance use disorders had lower completion odds, aligning with prior research highlighting barriers related to cognitive burden, engagement, and symptom severity (28). In contrast, patients with anxiety, ADHD, autism spectrum disorder (ASD), Cluster B personality disorders, insomnia, and OCD had higher completion odds. These trends may reflect condition-specific traits such as inflexible behaviors, heightened emotional states, or focus on the completeness of tasks (29). Notably, patients with insomnia had high completion odds despite a small sample size, which may reflect high awareness of and desire to report sleep-related impairments. Tailored strategies like brief symptom-specific measures, proxy reporting, or assisted completion may improve participation among patients with more severe impairments.

### 4.3 ePRO Implementation

We found that applying Learning Evaluation and FRAME frameworks enabled systematic feedback collection and responsive system modifications, aiding stakeholder communication. RE-AIM added clarity with concise, comprehensible measurements that communicate the real-world impact of an intervention.

#### 4.3.1 Implementation Facilitators

Multi-modal ePRO delivery (email, SMS, QR) improved accessibility, with SMS contributing significantly to increased reach. EMR integration automated deployment and reporting, reduced administrative burden, and facilitated real-time decision-making that was critical for provider acceptance (30). The brevity and clinical relevance of PROMIS tools minimized response burden and promoted clinician engagement, while Spanish-language options reduced language barriers.

Sustained staff engagement via governance meetings and targeted training helped address staff turnover and informed successful modifications such as “quick registration”. Approximately 9.5 hours were dedicated to ePRO-related meetings between August 2021 and December 2023, showing that meaningful engagement can be maintained with minimal time burden.

#### 4.3.2 Implementation Barriers

Barriers included technical issues and system usability concerns. These were mitigated through training and iterative improvements, underscoring the importance of a user-centered design approach and agile technical infrastructure. Technical issues during the SMS rollout initially caused workflow disruptions, highlighting the need for robust technical infrastructure and standardized contingency protocols. Early clinician skepticism, particularly around documentation burden and clinical relevancy, was addressed by incorporating their feedback, aligning ePROs with clinical relevance and CMS quality metrics—ultimately fostering acceptance.

### 4.4 Clinical Implications

The findings of this study can inform scalable ePRO implementation in other urgent care settings. Multi-modal delivery, brief validated measures, and EMR integration streamline data collection while minimizing burden. Structured feedback systems like governance meetings promote sustained engagement and continuous improvement. These strategies, grounded in implementation science and product development frameworks, are adaptable to a wide range of healthcare contexts requiring IT integration, enhancing interoperability, planning, testing, deployment, and training.

Future work should address disparities in completion, especially for patients with cognitive impairments, and further refine strategies to enhance the sustainability of ePROs with clinical, administrative, and institutional priorities. As healthcare moves toward value-based care, ePROs will play a critical role in capturing patient-centered outcomes and improving quality of care.

### 4.5 Limitations

Implementation strategies were tailored to the urgent care setting and may require adaptation for other clinical settings or patient populations. The study design lacked randomization, limiting causal inference, though a randomized trial was impractical due to workflow constraints.

## 5. Conclusion

This study demonstrates that ePROs can be effectively and sustainably implemented in psychiatric urgent care settings. Importantly, this work demonstrates that ePROs can be successfully integrated into fast-paced clinical environments. Key facilitators included multi-modal delivery, EMR integration, and continuous staff engagement through governance processes. Addressing disparities in ePRO completion and optimizing clinical utility are important next steps, and the frameworks used here offer a model for broader implementation in complex real-world healthcare environments.

## Declarations

### Ethics approval and consent to participate

This project was undertaken as a Quality Improvement/Evidence-Based Practice Initiative at Sheppard Pratt, and as such was not formally supervised by the Sheppard Pratt Institutional Review Board.

### Consent for publication

Not applicable.

### Availability of data and materials

The datasets used and/or analyzed during the current study are available from the corresponding author upon reasonable request.

## Authors’ contributions

AB managed and reviewed the pilot and implementation phases of the intervention, analyzed ePRO data and qualitative feedback, and was the lead contributor in writing the manuscript. EAB managed and reviewed the pilot and implementation phases of the intervention, assisted with reviewing ePRO data, and assisted with editing the manuscript. AR and AM provided data management and assisted with writing the “Methods” section of the manuscript. MP supervised data management. RS supervised all phases of the intervention and data analysis. All authors read and approved the manuscript.

## Acknowledgements

We thank the urgent care supervisor, Lynn Stefanowicz, for her assistance with scheduling ePRO meetings and supervising the urgent care staff on ePRO workflows. We thank the urgent care managers, Sharnice Coleman and Kristi Fleming, and their teams for their continuous feedback, efforts to encourage support for ePRO implementation, and communication with patients about ePROs. We thank the urgent care clinical supervisor, Dr. Laura Eskander, for providing feedback about clinically relevant ePROs and assisting with ePRO Adoption among urgent care clinicians.

