## Supplemental Tables and Figures for "Implementing electronic patient-reported outcome measures in psychiatric urgent care"

Supplementary Table 1. Description of ePRO implementation team

| ePRO Implementation Team Identifier | Point-of-care or technical | Professional Title | Degrees, Licenses, Certifications | ePRO duties |
|---|---|---|---|---|
| ePRO Director - Clinical | Point-of-care | Board certified psychiatrist | MD | Supervises clinical implementation and technology scalability |
| ePRO Director - Data Management | Technical | Director | Masters | Supervises data staging and reporting, project management, and interdepartmental communication |
| ePRO project coordinator | Point-of-care & Technical | Data Engineer & Project Coordinator | MHS | Monitors distribution and completion of ePROs, develops metric reporting, assists with user experience development/design, leads governance meetings and modification tracking |
| ePRO project coordinator | Point-of-care | Project Coordinator, Licensed Graduate Professional Counselor | MPH, LGPC | Monitors distribution and completion of ePROs, assists with user experience development/design, leads governance meetings and modification tracking |
| Senior Data Engineer | Technical | Data Engineer Sr | BS | Full stack development of ePRO system - system administration, technical design, systems integration |
| Senior Data Engineer | Technical | Data Engineer Sr | Masters | Full stack development of ePRO system - system development and design, user experience development/design |
| Data Engineer | Technical | Data Engineer | Masters | Maintain REDCap and ePRO dashboard interface to allow ePROs to be sent to patients |
| Data Engineer | Technical | Data Engineer | BS | Manage the distribution of ePROs and maintain integrated dashboards; providing technical support to ensure reliable workflows |
| Junior Data Engineer | Technical | Data Engineer Jr | BS | Address immediate user needs and requests and build out new dashboard concepts |

Supplementary Table 1 details the ePRO implementation team members. ePRO implementation Team Identifier refers to the name used throughout the study. Point-of-care or technical refers to whether the member worked directly with urgent care staff or if their work was restricted to developing and monitoring ePRO technology. ePRO duties refer to the specific ePRO tasks and activities performed.

Supplementary Table 2. Description of urgent care stakeholders

| ePRO Implementation Identifier | Clinical or Administrative | Professional Title | Degrees, Licenses, Certifications | ePRO duties |
|---|---|---|---|---|
| Lead Administrator | Administrative | Chief of Access & Integrated Care | MSW, LCSW | Supervise implementation of ePRO procedures; perform regular ePRO metric reviews with staff; attend governance meetings |
| Clinical Lead 1 | Clinical | Medical Director and Senior Psychiatrist | MD | Supervise clinical utility of ePRO data by reviewing EMR appointment notes for ePRO data; monitor ePRO documentation among clinicians; communicate ePRO updates to clinical staff; triage feedback to ePRO implementation team as needed; attend governance meetings |
| Clinical Lead 2 | Clinical | Psychiatrist | MD | |
| Administrator - Location T | Administrative | Manager | BA | Supervise administrative procedures for ePROs including “quick registration” and dashboard monitoring; training administrative staff on communicating with patients about ePROs; triage feedback to ePRO implementation team as needed; attend governance meetings |
| Administrator - Location B | Administrative | Manager | BA | |

Supplementary Table 2 details the urgent care site governors and their professional experience. ePRO implementation Identifier refers to how the individual is referred throughout the study. Clinical or administrative refers to whether the team member completed clinical (i.e. treat patients) or administrative (i.e. register patients) duties. ePRO duties refer to the specific ePRO tasks and activities performed by the site governor.

Supplementary Figure 1. ePRO workflow

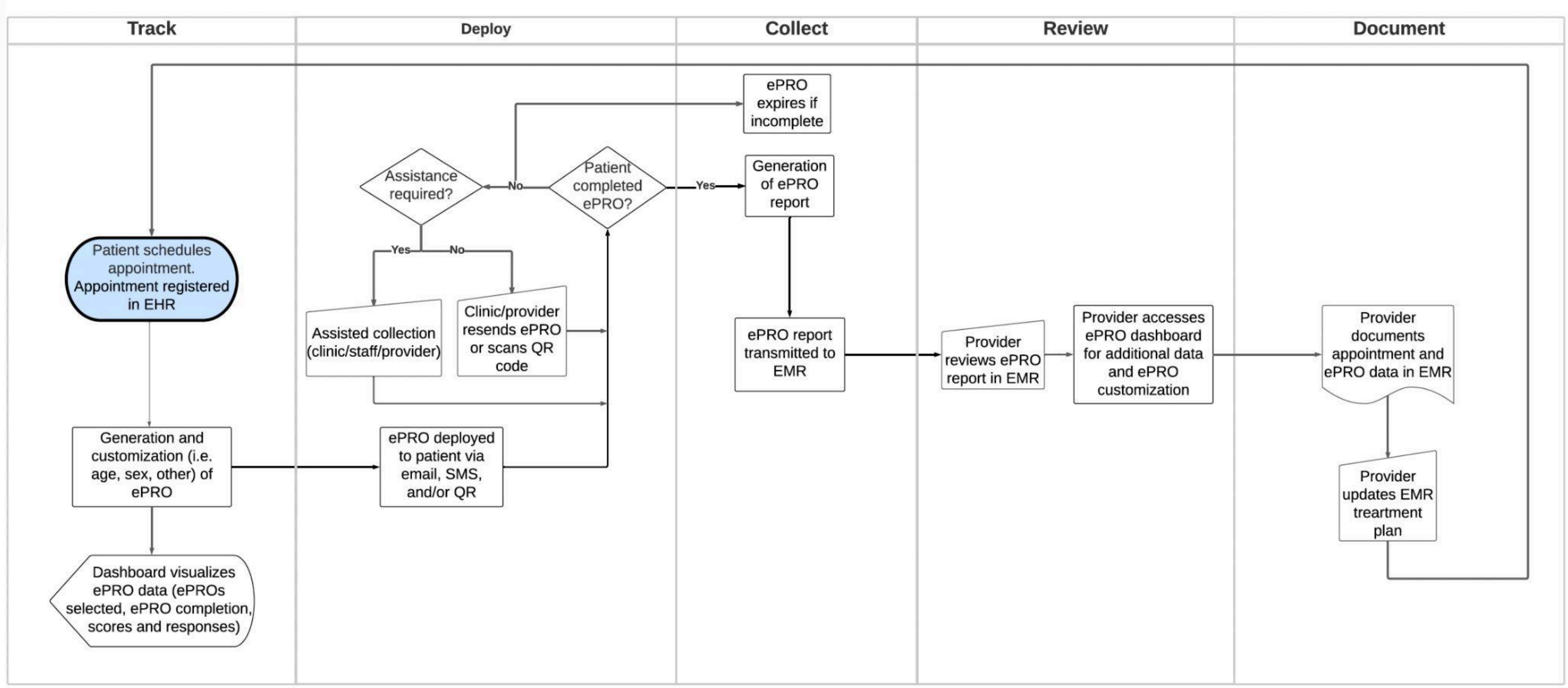


Supplementary Figure 1 shows the ePRO workflow based on an implementation guide by LeRouge et al.. The figure displays how the ePRO system handles ePROs for patients, administrative staff, and clinical staff. The descriptions of each workflow component are:

Deploy - sending ePROs to the patient
Collect - ePRO completion
Track - monitoring of ePRO completion
Review - accessing and viewing the ePRO data
Document - saving ePRO data over time

Supplementary Figure 2a. Example of ePRO deployment

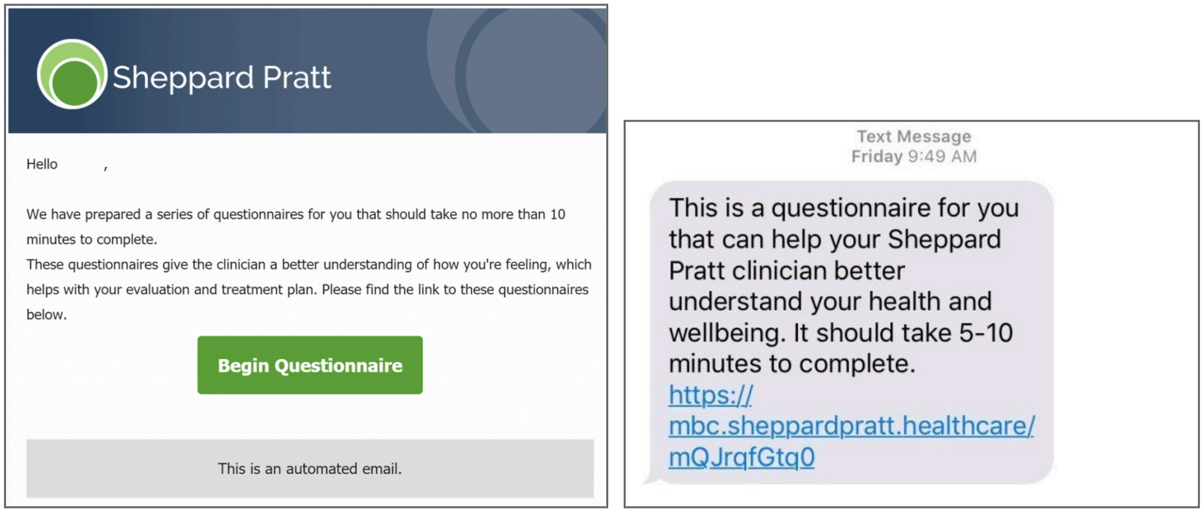


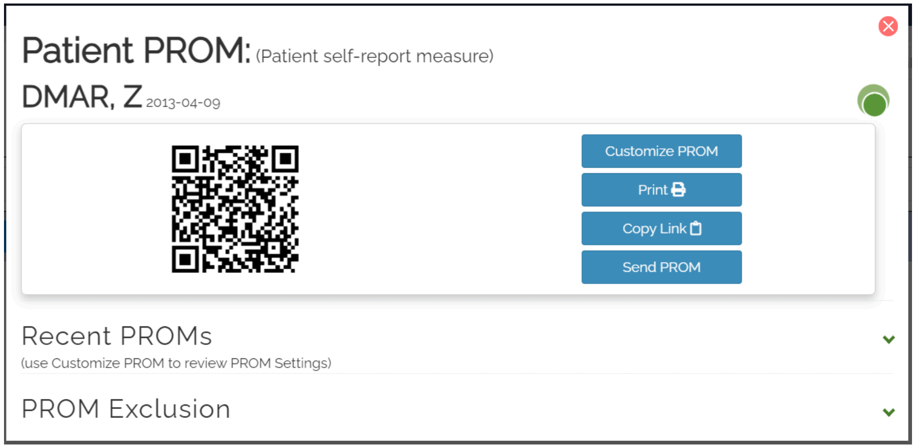


Supplementary Figure 2b. Example of translation selector

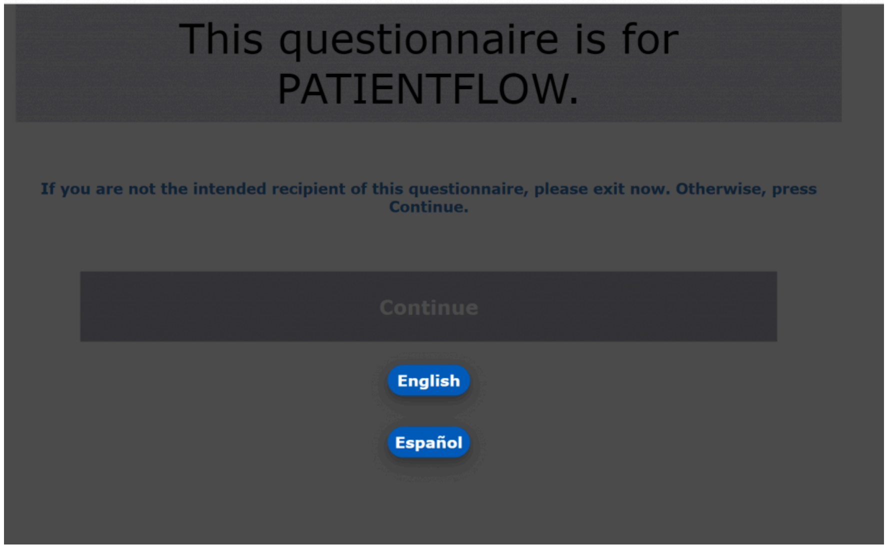


Supplementary Figure 2a displays the ePRO email (left), ePRO SMS/text (right) notifications that patients receive for automated and on-demand ePRO deployment (if there is a patient email and/or cell phone number entered in the EHR). The text used in the email and SMS/text notification is standardized for all Sheppard Pratt patients, however, the links are unique for each patient and ePRO instance.

The third figure for Supplementary Figure 2a shows the user interface (UI) for ePRO deployment on the ePRO dashboard. The QR code, when scanned with a smart device, opens to the current ePROs that are assigned to the patient. The QR code can be printed using “Print”. The link associated with the QR code can also be copied using “Copy Link” or sent to the patient’s email or cell phone number using “Send PROM”. Additional features are available including displaying the patient’s recent ePRO completion history (“Recent PROMs"), displaying reasons for exclusion from ePROs (“PROM Exclusion”), and adding or removing ePROs (“Customize PROM”).

Supplementary Figure 2b shows the UI for patients upon opening their ePRO link. The screen displays language options for English and Spanish. Once selected, the language options move to the top corner of the screen, allowing patients the option to change the language as needed as they complete the ePRO.

Supplementary Table 3. ePRO battery

| Measure | Age Range | Was the measure modified? | If yes, how was the measure modified? |
|---|---|---|---|
| PROMIS Short Form v1.0 - Depression 4a | Adult (18+) | No | |
| PROMIS Pediatric Short Form v2.0 - Depressive Symptoms 8a | Pediatric (12-17) | No | |
| PROMIS Parent Proxy Short Form GenPop v2.0 - Depressive Symptoms 6a | Proxy (5-11) | No | |
| PROMIS Short Form v1.0 - Anxiety 4a | Adult (18+) | No | |
| PROMIS Pediatric Short Form v2.0 - Anxiety 8a | Pediatric (12-17) | No | |
| PROMIS Parent Proxy Short Form v2.0 - Anxiety 8a | Proxy (5-11) | No | |
| PROMIS Short Form v1.0 - Sleep Disturbance 4a | Adult (18+) | No | |
| PROMIS Pediatric Short Form v1.0 - Sleep Disturbance 4a | Pediatric (12-17) | No | |
| PROMIS Parent Proxy Short Form v1.0 - Sleep Disturbance 4a | Proxy (5-11) | No | |
| PROMIS Short Form v1.0 - Sleep-Related Impairment 8a | Adult (18+) | Yes | Patient only completed this measure if they scored as moderate or higher on PROMIS Sleep Disturbance |
| PROMIS Pediatric Short Form | Pediatric (12-17) | Yes | Patient only completed this |

| | | | |
|---|---|---|---|
| v1.0 - Sleep-Related Impairment 4a | | | measure if they scored as moderate or higher on PROMIS Sleep Disturbance |
| PROMIS Parent Proxy Short Form v1.0 - Sleep-Related Impairment 4a | Proxy (5-11) | Yes | Patient only completed this measure if they scored as moderate or higher on PROMIS Sleep Disturbance |
| AUDIT-C | Pediatric (12-17),<br>Adult (18+) | No | |
| Proxy AUDIT-C | Proxy (5-11) | Yes | Modified for parent proxy reporting. |
| PROMIS Short Form v1.0 - Alcohol Use 7a | Pediatric (12-17),<br>Adult (18+) | Yes | Patient only completed this measure if the AUDIT-C was positive |
| PROMIS Proxy-Edited Short Form v1.0 - Alcohol Use 7a | Proxy (5-11) | Yes | Modified for parent proxy reporting. This was only completed if the AUDIT-C was positive. |
| PROMIS Short Form v1.0 - Severity of Substance Use (Past 30 days) 7a | Pediatric (12-17),<br>Adult (18+) | Yes | The patient only completed this measure if they indicated using illicit, non-prescribed drugs on the NIDA screener. |
| PROMIS Proxy Short Form v1.0 - Severity of Substance Use (Past 30 days) 7a | Proxy (5-11) | Yes | Modified for parent proxy reporting. The patient only completed this measure if they indicated using illicit, non-prescribed drugs on the screener question. |
| PROMIS Short Form v2.0 - Physical Function 4a | Adult (18+) | No | |

| | | | |
|---|---|---|---|
| PROMIS Pediatric Short Form v2.0 – Mobility 8a | Pediatric (12-17) | No | |
| PROMIS Parent Proxy Short Form v2.0 – Mobility 8a | Proxy (5-11) | No | |
| Tobacco Use Screen | Pediatric (12-17), Adult (18+) | Yes | Created in collaboration with the EHR team and urgent care clinical managers. |
| Fall Screen | Geriatric (65+) | Yes | Created in collaboration with the EHR team and urgent care clinical managers. |
| Vulnerability to Abuse Screening Scale (VASS) | Geriatric (65+) | No | |

Supplementary Table 3 displays the psychometric instruments selected for the ePRO battery used for the intervention. These instruments were selected based on their clinical relevance for the urgent care patient population. Some instruments were modified, such as the modification of adult versions to proxy-report versions, based on the clinical interest of the urgent care clinicians.

Supplementary Figure 3. Governance meeting process

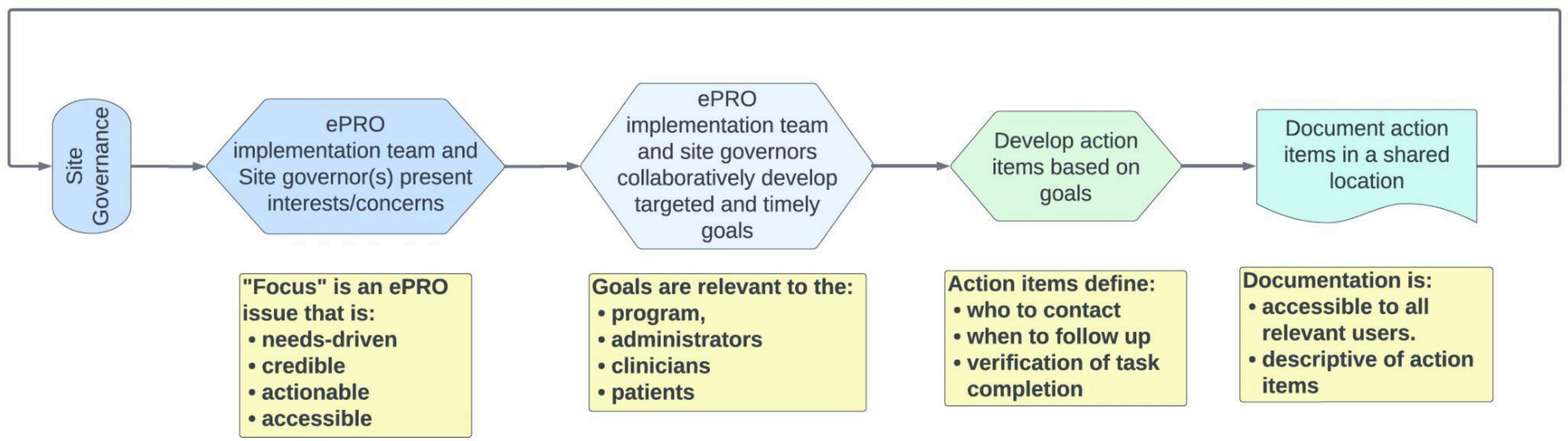


Supplementary Figure 3 displays the site governance process used for ePRO implementation. Site governance meetings occur monthly and are restricted to key stakeholders for a program or care level. If needed, the stakeholders can schedule ePRO trainings for their staff with ePRO project coordinators as an action item.

The governance meeting process was developed based on the components of knowledge transfer and exchange by Prihodova et al. The purpose of this process is to identify key items affecting ePRO implementation (Focus), develop achievable goals to address the key items (Goals), develop procedures to achieve the goals (Action items), and document the focus, goals and action items (Documentation). This process allows key stakeholders and staff to be engaged and accountable for ePRO implementation, along with the ePRO implementation team.

Supplementary Table 4. Diagnostic categories and associated ICD codes

| **Diagnostic Category** | **ICD Codes** |
|---|---|
| ADHD | F79, F90.9, F90.2 |
| Adjustment Disorders | F43.2 |
| Alcohol and Substance Use Disorders | F10.1, F10.2, F10.9, F11, F11.1, F11.2, F11.9, F12.1, F12.2, F12.9, F13, F13.2, F14, F14.1, F14.2, F15, F19, F19.1, F19.9, F40, F63 |
| Anxiety Disorders | F40, F40.1, F41, F41.0, F41.1, F41.8, F41.9 |
| ASD | F84.0 |
| Bipolar Disorders | F30, F31, F31.2, F31.3, F31.4, F31.6, F31.7, F31.8, F31.9 |
| Cluster B Personality Disorders | F60, F60.3, F60.9 |
| Depressive Disorders | F32, F32.A, F32.2, F32.3, F32.8, F32.9, F33, F33.0, F33.1, F33.3, F33.4, F33.9, F34, F53 |
| DMDD | F63.9 |
| Eating Disorders | F50, F50.8, F50.9 |
| Gender Dysphoria | F64 |
| Impulse Control Disorders | F63.8, F91 |
| Insomnia | G47, G47.0 |
| Neurocognitive Disorders | F01, F03, F07, F09, F84, G30, G31.8, R41, S06, Z87 |
| Neurodevelopmental Disorders, Other | F70, F71, F72, F79, F81, F89, F95 |
| OCD | F42, F42.9 |
| Psychosis, Schizophrenia Spectrum, Delusional Disorders | F06, F17.2, F20, F20.9, F22, F23, F25, F25.9, F29 |
| Severe and Treatment Resistant Mood Disorders | F34.1, F39 |
| Somatic Disorders | F44, F45 |
| Trauma Disorders | F43, F43.1, F44.8 |

Supplementary Table 4 displays the ICD codes extracted from the EMR for urgent care patients, grouped into clinically relevant diagnostic categories based on DSM-5 chapters.

Supplementary Table 5. RE-AIM dimensions applied to ePRO metrics

| RE-AIM Dimension | Dimension Definition | ePRO Implementation Measure |
|---|---|---|
| Reach | The number, proportion, or representativeness of admitted patients completing ePROs | ePRO completion<br><br>(Number of patients with a completed ePRO / the number of patients admitted) |
| Adoption | The number, proportion, or representativeness of clinicians documenting ePRO data | ePRO documentation<br><br>(Number of patients with an ePRO populated in an EMR appointment note / the number of patients with completed ePROs) |
| Implementation | The identification, time, and usage of intervention protocols and strategies | Number of meetings with urgent care staff |
| | | Number of ePRO system modifications |
| | | Time to complete ePRO system modifications |
| Maintenance | The extent to which the ePRO implementation strategies become part of urgent care practices | ePRO completion remains above average |
| | | Active use of ePRO modifications |

Supplementary Table 5 displays how RE-AIM dimensions were aligned with the ePRO intervention metrics. Each ePRO outcome measure was analyzed quantitatively and qualitatively.

Supplementary Table 6. ePRO modifications

| Modification ID for Figure 1 | Modification description | Modification owner | Time from request to implementation | Completion date | Story Points |
|---|---|---|---|---|---|
| Mod #1 | Add Location B to the ePRO dashboard | ePRO implementation team | 2 days | August 2, 2021 | 1 |
| Mod #2 | Add substance use screener to ePRO battery | ePRO implementation team | 4.5 months (146 days) | March 22, 2022 | 2 |
| Mod #3 | Add location filter on ePRO dashboard | ePRO implementation team | 4 months (121 days) | August 19, 2022 | 2 |
| Mod #4 | Add ePROs to the quality metrics tracking system | ePRO implementation team and EHR department | 2 months (73 days) | October 27, 2022 | 3 |
| Mod #5 | Add SMS option for sending ePROs to patients | ePRO implementation team | 5 months (156 days) | May 5, 2023 | 6 |
| Mod #6 | Use "quick registration" workflow for all patients | Urgent care staff | Location B – approximately 8 months (256 days)<br><br>Location T - approximately 11 months (344 days) | Location B - no specific date defined; staff provided confirmation of full time usage June 15, 2023<br><br>Location T - no specific date defined; staff was instructed on September 11, 2023 to use workflow full time | 3 - Although urgent care staff did not utilize story points to calculate level of effort, the estimation is based on the level of effort of the ePRO implementation team for coordination and follow up with the urgent care staff. |

Supplementary Table 6 displays the modifications implemented throughout the ePRO implementation period. The "Modification owner" is the individual or group that was in charge of completing the modification. "Time from request to implementation" refers to the amount of time needed to build, test, and release the completed modification. "Completion date" refers to the exact date that the modification was completed and presented to users or when users confirmed that the modification was complete. Mod #6 had two release dates since the completion was confirmed by the individual urgent care locations at different times. “Story Points (SP)” refers to the units of measure for the level of effort required to complete the modification by the ePRO implementation team. The story points reflected in this column were tracked on user story cards completed during the sprints. The minimum story point value is 1, indicating the task was lower in complexity and amount of work to complete. Increased complexity, risk to functionality, and amount of work results in story point increases based on the Fibonacci sequence (1, 2, 3, 5, 8, etc.). Some modifications were completed across multiple user story cards thus the story points reflected in this table reflect the total story points across multiple cards.

Supplementary Table 7. Examples of common themes found in qualitative feedback for ePRO implementation

| **Theme** | **Category** | **Example comment** |
|---|---|---|
| Clinical Adoption | Barrier | "[ePRO] is not helpful for us. This is a very fast paced clinic. This is one more step [for us to complete]. This is defensive medicine, and takes effort because we need to then justify why a patient is not 'severe' if they test severe. This is a hurdle." |
| Administrative Process, Time | Barrier | "Unless there is paperwork to review, the patient is 'arrived' pretty quickly and there isn't much time for the patient to check the email to complete [ePRO]." |
| Administrative Process | Facilitator | "We started using a 'quick registration' process, where we have patients provide their name, [cell], and email as soon as they come in. This helps us to quickly get their info into [the EHR] while they're completing the rest of the registration packet. " |
| Administrative Process, Time | Barrier | "For [Location T], the [quick registration] usage is probably low because when a provider is ready to take a patient, we go ahead and assign the patient to that provider since wait times tend to be low." |
| Clinical Adoption, Time | Facilitator | "It's better for the provider to have ePROs completed [before seeing the patient]. Some of those [quality] requirements are met…It reduces workload for the provider." |
| Administrative Process | Facilitator | "Using the quick registration sheet helps with getting PROMs completed. We've been using it for a while now…We also give reminders to patients who are still in the waiting room who haven't completed it." |
| Administrative Process | Facilitator | "Our staff here work really well with the patients and are very communicative. [Administrator] has been great with getting the admins trained, including the weekend and evening teams. I think, as a team, we're all on the same page with how we're handling PROM so there's no confusion about what to do or how to handle any patient questions." |
| Administrative Process | Facilitator | "The quick registration process has been working. We've noticed that the patients are receiving the links faster and are able to complete it as they're finishing their paperwork. It also helps us with assigning them to a provider. It's nice to see that there are improvements in the metrics." |

Supplementary Table 7 displays examples of the qualitative feedback collected throughout the ePRO intervention. Deductive and inductive coding were used to determine the "Theme". "Category" identifies if the comment refers to helpful (facilitator) or unhelpful (barriers) aspects of ePRO implementation.

Supplementary Table 8. ePROs used for CMS Quality Payment Program program Quality Metrics

| Quality ID | Name | Description | ePROs used |
|---|---|---|---|
| 134 | Preventive Care and Screening: Screening for Depression and Follow-Up Plan | Percentage of patients aged 12 years and older screened for depression on the date of the encounter or up to 14 days prior to the date of the encounter using an age-appropriate standardized depression screening tool AND if positive, a follow-up plan is documented on the date of the eligible encounter. | PROMIS Short Form v1.0 - Depression 4a<br><br>PROMIS Pediatric Short Form v2.0 - Depressive Symptoms 8a |
| 226 (NQF 0028) | Preventive Care and Screening: Tobacco Use: Screening and Cessation Intervention | Percentage of patients aged 18 years and older who were screened for tobacco use one or more times within the measurement period AND who received tobacco cessation intervention on the date of the encounter or within the previous 12 months if identified as a tobacco user | Tobacco Use Screen |
| 402 | Tobacco Use and Help with Quitting Among Adolescents | The percentage of adolescents 12 to 20 years of age with a primary care visit during the measurement year for whom tobacco use status was documented and received help with quitting if identified as a tobacco user | Tobacco Use Screen |
| 431 (NQF 2152) | Preventive Care and Screening: Unhealthy Alcohol Use: Screening &<br>Brief Counseling | Percentage of patients aged 18 years and older who were screened for unhealthy alcohol use using a systematic screening method at least once within the last 12 months AND who received brief counseling if identified as an unhealthy alcohol user | AUDIT-C |
| 182 (NQF 2624) | Functional Outcome Assessment | Percentage of visits for patients aged 18 years and older with documentation of a current functional outcome assessment using a standardized functional outcome assessment tool on the date of the encounter AND documentation of a care plan based on identified functional outcome deficiencies on the date of the identified deficiencies | PROMIS Short Form v2.0 - Physical Function 4a |
| 155 (NQF | Falls: Plan of Care | Percentage of patients aged 65 | Fall Screen |

| | | | |
|---|---|---|---|
| 0101) | | years and older with a history of falls that had a plan of care for falls documented within 12 months | |
| 181 | Elder Maltreatment Screen and Follow-Up Plan | Percentage of patients aged 65 years and older with a documented elder maltreatment screen using an Elder Maltreatment Screening tool on the date of encounter AND a documented follow-up plan on the date of the positive screen. | Vulnerability to Abuse Screening Scale (VASS) |
| PP10 | Measurement-Based Care Processes: Index Assessment, Monitoring and Care Plan Review | Percentage of individuals 18 years of age and older with a mental and/or substance use disorder, who had a comprehensive index assessment in the measurement period, with monitoring and care plan review. | PROMIS Short Form v1.0 - Depression 4a<br><br>PROMIS Short Form v1.0 - Anxiety 4a<br><br>PROMIS Short Form v1.0 - Severity of Substance Use (Past 30 days) 7a<br><br>AUDIT-C |

Supplementary Table 8 displays the CMS Quality Payment Program program Quality Metrics utilized by urgent care and the ePROs that were used to complete the metric. The full names of and descriptions of the quality metrics are listed. Some metrics were associated with multiple ePROs based on the age of the patient or the comprehensive nature of the metric.
